# How to choose a time zero for patients in external control arms

**DOI:** 10.1101/2020.12.02.20242040

**Authors:** Daniel Backenroth

## Abstract

When a sponsor carries out a single-arm trial of a novel oncology compound, it may wish to assess the efficacy of the compound via comparison of overall survival to an external control arm, constructed using patients included in some retrospective registry. If efficacy of the novel compound is compared to efficacy of physician’s choice of chemotherapy, patients in the retrospective registry might qualify for inclusion in the external control arm at multiple different points in time, when they receive different chemotherapy treatments. For example, a patient might qualify at the start of their second, third and fourth lines of therapy. From the start of which line of therapy should this patient’s survival be compared to survival of participants in the single-arm trial?

Some sponsors have elected to include patients in the external control arm from the last available line of therapy in the retrospective database. Another possibility is to randomly select a line of therapy for each external control arm patient from among those available. In this paper, we show, via probabilistic arguments and also via simulation based on real data, that both of these methods give rise to a bias in favor of the single-arm trial. We further show that this bias can be avoided by instead including external control arm patients multiple times in the external control arm, once for each time they receive qualifying treatment.

## Introduction

Imagine that a sponsor is carrying out a single-arm trial of a novel oncology compound for patients with advanced non-small cell lung cancer. Since this is the first clinical trial for the compound, heavily pre-treated participants have been enrolled. In particular, participants are required to have been treated with prior platinum chemotherapy. Initial regulatory approval of novel oncology compounds could be based on such a single-arm trial [1, 2]. However, to provide additional context to regulators, the sponsor has decided to construct an external control arm for the trial, using data retrospectively collected from an electronic health records system [3, 4]. The sponsor will compare overall survival for patients in the external control arm to overall survival for patients in the single-arm trial, after reweighting to account for differences in patient characteristics.

One of the first steps in constructing an external control arm is to apply appropriate inclusion and exclusion requirements to the retrospective database. Here, one such inclusion criteria might be that the patient has started physician’s choice of anticancer treatment after previous treatment with platinum chemotherapy [5]. Survival from the start of this anticancer treatment for patients in the external control arm will be compared to survival from receipt of the first dose of the novel oncology compound in the single-arm trial.

One problem in applying this criterion is that the sponsor may very well find that some patients in the retrospective database satisfy it at multiple different points in time [6, 7]. For example, consider the two patients in Table 1. Patient P2 has only one line of therapy (docetaxel) after their platinum line, and so would only qualify for inclusion in the external control arm with respect to that line of therapy. On the other hand, Patient P1 has two lines of therapy (first docetaxel plus ramucirumab, and then pemetrexed) after their platinum line, and so qualifies for inclusion in the external control arm twice. From what “time zero” should overall survival be measured for patient P1 in the external control arm: the beginning of their second or the beginning of their third line of therapy? The timepoint selected may have a large effect on the results of the comparison to the single-arm trial, since a patient clearly survives longer from an earlier line of therapy than from a later line of therapy. Patient P1, for example, survived for nearly a year from the start of second-line treatment with docetaxel plus ramucirumab, but for less than three months from the start of third-line treatment with pemetrexed.

**Table 1:**
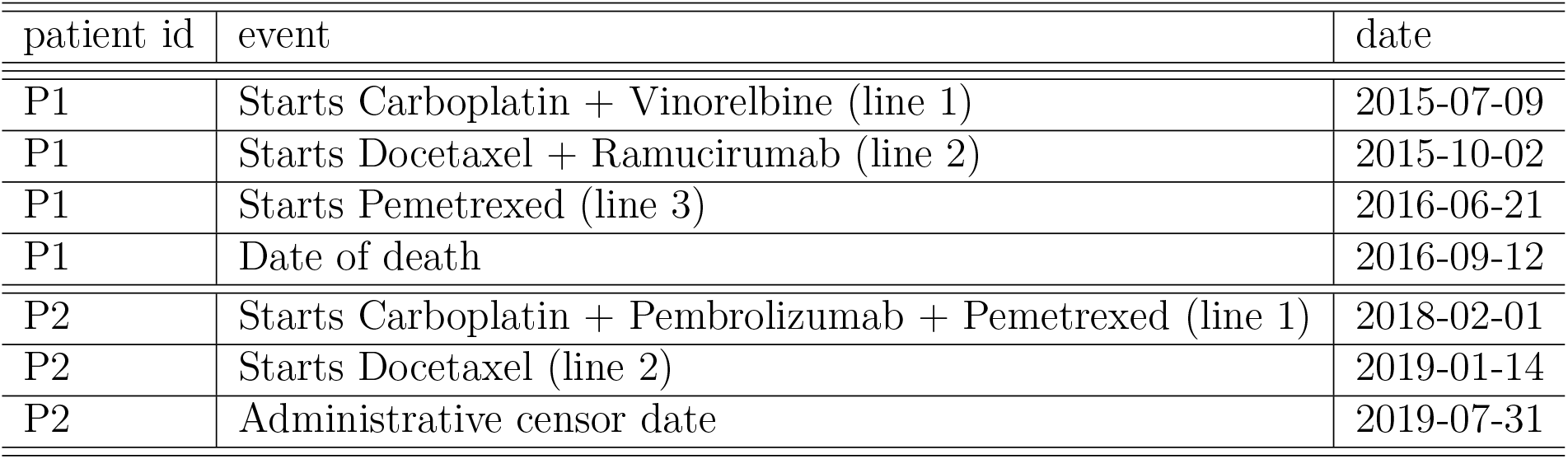
Treatment history and known vital status for two fictitious patients in a retrospective database

| patient id | event | date |
| --- | --- | --- |
| P1 | Starts Carboplatin + Vinorelbine (line 1) | 2015-07-09 |
| P1 | Starts Docetaxel + Ramucirumab (line 2) | 2015-10-02 |
| P1 | Starts Pemetrexed (line 3) | 2016-06-21 |
| P1 | Date of death | 2016-09-12 |
| P2 | Starts Carboplatin + Pembrolizumab + Pemetrexed (line 1) | 2018-02-01 |
| P2 | Starts Docetaxel (line 2) | 2019-01-14 |
| P2 | Administrative censor date | 2019-07-31 |

Hernán and Robins have discussed the problem of patients in retrospective databases with multiple eligible time zeros, in the context of studies that compare two groups both extracted from the same retrospective database. They suggested that for patients with multiple eligible time zeros, there were two unbiased approaches [8]. A single eligible time, like the first eligible time or a random time, could be chosen as the time zero for each patient. Alternatively, all eligible times could be used, so that different versions of the same patient would be included in the comparison, each with a different time zero.

In this article, we describe a general model of cancer patient survival, of enrollment in clinical trials and of inclusion in retrospective database, and show that several methods of choosing a single time zero, like choosing the start of the last available line of therapy as in [6, 7], or choosing a randomly selected line of therapy, are not unbiased for an overall survival endpoint when, unlike in Hernán and Robins’ setting, comparison is made between a retrospective database and a clinical trial (as opposed to solely within a retrospective database). We then demonstrate how the other method suggested by Hernán and Robins, in which the same patient is included multiple times in the same comparison, but with different time zeros, can be used to obtain an unbiased comparison of overall survival between a single-arm trial and patients in a retrospective database.

## Model

Here we describe a general model of the survival of cancer patients, of how they enroll in a clinical trial and of how they are included in a retrospective database. As above, we take as an example population of interest non-small cell lung cancer patients who have received platinum chemotherapy and go on to receive subsequent therapy. We will see how survival of patients enrolled in a hypothetical control arm of a trial in which patients are treated with physician’s choice of standard of care therapy compares to the survival of patients in an external control arm treated with physician’s choice of standard of care therapy, given various choices of time zero for external control arm patients. We should choose a time zero for external control arm patients such that the survival distribution of patients in the external control arm is the same as the survival distribution of patients in this hypothetical control arm, assuming all other requirements for causal inference, e.g., no unmeasured confounders, are satisfied [9]. We call the trial control arm hypothetical since our focus here is on single-arm trials, in which there is no control arm.

Assume that each patient *i* in the population of interest has a time *S*_*i*_, measured from some arbitrary common timepoint, for example, January 1, 2000, at which they start their first post-platinum therapy, and has *N*_*i*_ post-platinum lines of therapy before they die, where *N*_*i*_ *≥* 1. The length of patient *i*’s *j*th post-platinum line of therapy is *D*_*ij*_. We assume that patients die immediately after the end of their *N*_*i*_th line of therapy. We further assume that the *D*_*ij*_ are independent of *S*_*i*_, so that there is no time trend in survival, but that the *D*_*ij*_ can depend on *N*_*i*_, so that patients with more lines of therapy can have longer (or shorter) lines of therapy. Conditional on *N*_*i*_, however, we assume that *D*_*ij*_ is independent of *D*_*ik*_ for all *j ≠ k*. Since we are focusing on patients receiving standard of care therapies, we assume that *D*_*ij*_ and *D*_*ik*_, for *k > j*, do not depend on whether patient is enrolled in a clinical trial in line *j*. Due to censoring, *N*_*i*_ and some of the *D*_*ij*_ may not be observed for some patients.

### Clinical trial

We assume that a trial enrolls participants from this population between times *t*_1_ and *t*_2_, measured from the same common baseline time as the *S*_*i*_, and that every patient in the population that finishes first-line platinum therapy during this time period, or that finishes a subsequent line of therapy during this time period and that doesn’t die at the end of that line of therapy, enrolls in the trial with probability *α*.

Let *E*_*i*_ be independent random variables that are each equal to 1 with probability *α* and equal to 0 with probability 1 − *α*. Under the model above patient *i* will enroll in the trial if *E*_*i*_ = 1 and if there exists a *k* in 0 … *N*_*i*_ − 1 such that

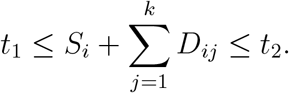

The sum in this expression equals 0 if *k* = 0, in which case the patient enrolls in the trial at time *S*_*i*_, as their first line of therapy after platinum therapy. The line of therapy, counting starting with the first post-platinum therapy, in which patient *i* will enroll in the trial is

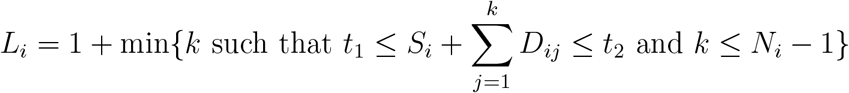

The survival of patient *i* from enrollment in the trial until death is 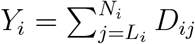. For some patients *Y*_*i*_ may be censored and not known at the time of comparison to the external control arm.

### Retrospective registry

We assume that a retrospective registry of cancer patients includes, with probability *β*, any patient from this population for whom *S*_*i*_ *< t*_*c*_, where *t*_*c*_ is the data cutoff date for the registry. That is, if the patient starts their line of treatment after platinum chemotherapy after the date *t*_*c*_, then they won’t be included in the registry. Let *F*_*i*_ be independent random variables that are each equal to 1 with probability *β* and equal to 0 with probability 1−*β*. We assume that the registry includes complete information until time *t*_*c*_ on the treatment history of any patient with *F*_*i*_ = 1 and *S*_*i*_ *< t*_*c*_. Any events occurring after time *t*_*c*_ are censored. The number of post-platinum lines of therapy observed for a patient in the registry depends on the relationship between *S*_*i*_, *t*_*c*_, *N*_*i*_ and the *D*_*ij*_. Letting *M*_*i*_ be the number of post-platinum lines of therapy whose start time is observed for patient *i* in the retrospective database, which may be lower than *N*_*i*_ due to censoring, we have that

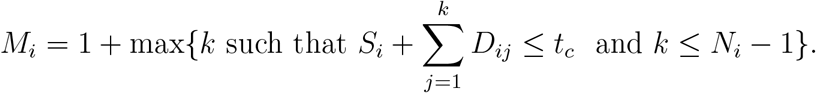

The survival of patient *i* from the start of their *k*th post-platinum line, 1 ≤ *k* ≤ *N*_*i*_, is 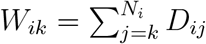, although this survival may be censored.

### Overall survival observed in clinical trial

We want to check if *P* (*Y*_*i*_ *≥ y*|*L*_*i*_ = *l, E*_*i*_ = 1), the survival function for patients enrolling in the trial in line setting *l*, is the same as 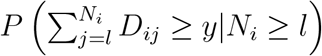, the marginal survival function for patients in the population, from that line setting, regardless of enrollment in the trial. We have

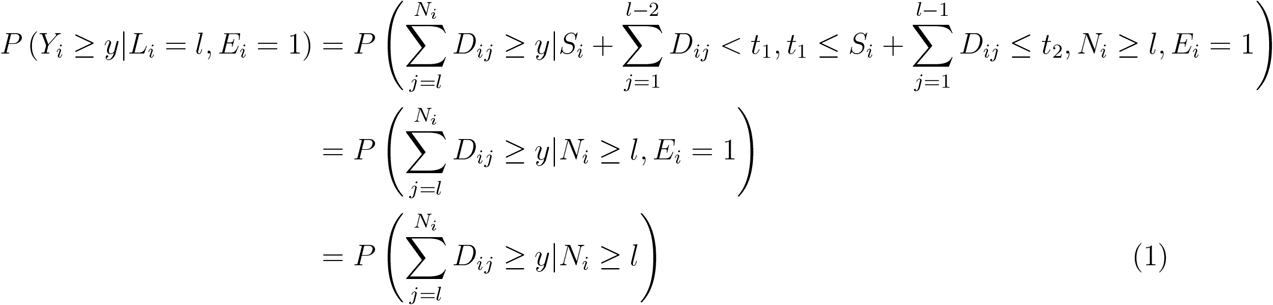

The first line above follows since a patient will enroll in the trial in line setting *l* only if the patient’s *l* − 1th post-platinum line is the first line of that patient to end in the interval between *t*_1_ and *t*_2_ (if *l* = 1 then the first inequality conditioned on is ignored). The second line follows from mutual independence of the *D*_*ij*_ and from mutual independence of the *D*_*ij*_ and the *S*_*i*_, and the third line follows from independence of the *D*_*ij*_ and *E*_*i*_. Therefore, under the model posed above, the survival of patients enrolling in the trial in a particular line setting *l*, i.e., those with *L*_*i*_ = *l*, is the same as the marginal survival of patients in the population from the *l*th line.

Conditioning on *N*_*i*_ = *n*, we can also write this marginal probability as follows:

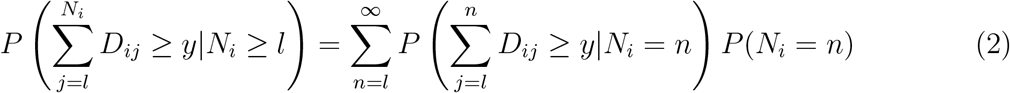

Note that equations 1 and 2 would not hold if the *D*_*ij*_ were not conditionally independent given *N*_*i*_. Here we sum to infinity, although in practice the *N*_*i*_ are bounded. In the appendix, we show, assuming that the times at which patients start post-platinum therapy *S*_*i*_ are uniformly distributed, that if the lengths of the *D*_*ij*_ are positively correlated within a patient then patients with better survival may be over-represented in a clinical trial.

### Overall survival observed in retrospective database, choosing one eligible line per patient

One potential way to set up an external control arm is to measure survival for each patient in the retrospective database from one post-platinum line of therapy chosen using some rule from the available post-platinum lines of therapy for that patient. Let *R*_*i*_ be the selected post-platinum line of therapy from which survival is measured for patient *i*. The selection of *R*_*i*_ could depend on *M*_*i*_. For example, one line might be chosen randomly from among the available lines of therapy in a database for a patient, or the last available line of therapy might be chosen for a patient. Alternatively, the selection of *R*_*i*_ could be independent of *M*_*i*_. For example, the first line could always be chosen, *R*_1_ = 1. As another example, *R*_*i*_ could equal 1 with probability 0.5 and 2 with probability 0.5, irrespective of *M*_*i*_. With this scheme, a patient with *R*_*i*_ *> M*_*i*_ would be excluded from the external control arm, and so sample size would be affected.

We first assume that we have complete capture for each patient in the retrospective registry, i.e., *N*_*i*_ is observed for each patient in the registry, so that *N*_*i*_ = *M*_*i*_. The survival for patients from line setting *l* in an external control arm so constructed, *P* (*W*_*il*_ *≥ y*|*R*_*i*_ = *l*), is, conditioning on *N*_*i*_ and then applying Bayes’ rule:

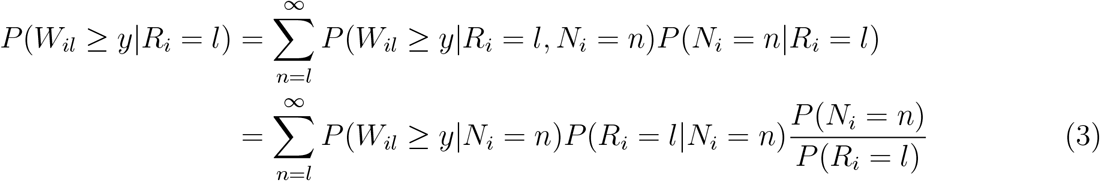

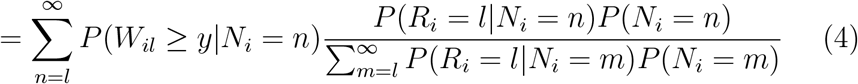

Here we sum *N*_*i*_ from *l* to *∞*, but the *N*_*i*_ in practice are bounded above. Note that in the second line we assume that *W*_*il*_ is independent of *R*_*i*_ given *N*_*i*_, i.e, that the selection of *R*_*i*_ may depend on how many lines of therapy a patient has, but not on other features of the survival history of the patient.

Let us consider some rules for selecting a post-platinum line of therapy for each patient in the external control arm. First we assume that we select a line uniformly at random for each patient from the available lines of therapy, that is, *P* (*R*_*i*_ = *l*| *N*_*i*_ = *n*) for *l*≤ *n* equals 1*/n*. Then equation 4 equals

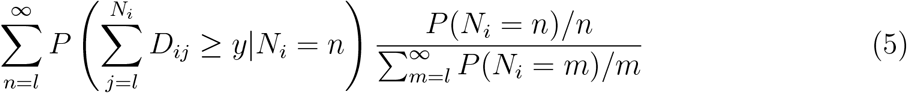

so we see that, by comparison to the weighted sum for the marginal survival probability observed in the trial (equation 2), probabilities for patients with more lines of therapy, and hence longer survival, are given smaller weights in this expression, leading to bias if this method is adopted.

In fact, by inspecting equations 2 and 3, we find that in general, in order to avoid bias, the selection of *R*_*i*_ for each patient must not depend on *N*_*i*_. So, for example, choosing the last line for each external control arm will lead to bias. In this case *P* (*R*_*i*_ = *l*|*N*_*i*_ = *n*) = 1 for *l* = *n* and 0 otherwise, so that equation 3 becomes *P* (*W*_*il*_*≥ y*| *N*_*i*_ = *l*), and we are only estimating survival from line setting *n* using patients who died immediately after that line setting.

By contrast, always choosing the first line for each patient, *P* (*R*_*i*_ = 1) = 1, will not lead to bias, but then no external control arm patients will be able to be compared to trial patients who received active comparator in a line after their first post-platinum line of therapy. Choosing a line of therapy at random for each patient where the random selection does not depend on *N*_*i*_ will avoid bias, but this choice will mean that many patients will be excluded from the external control if they did not survive until the randomly chosen line of therapy.

Before, we ignored the fact that we did not collect data for patients in the registry after the data cutoff date *t*_*c*_. Now we take this into account. First, we consider selection rules where *R*_*i*_ is independent of *M*_*i*_ and of the survival history of the patient. For example, assume we let *R*_*i*_ = 1 with probability 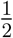 and *R*_*i*_ = 2 with probability 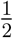. We want to check that *P* (*W*_*il*_ *≥ y*|*R*_*i*_ = *l, M*_*i*_ *≥ l, F*_*i*_ = 1),the survival of external control arm patients selected from the registry, is the same as 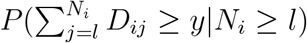, the marginal survival of patients from that line setting. If so, then we can use such selection rules to select an external control arm whose survival distribution can be compared without bias to the survival distribution of patients in the trial.

We have:

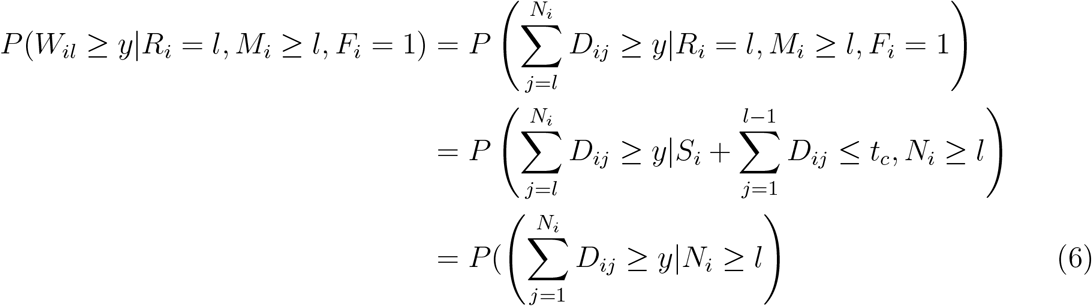

The second line here follows from the independence of *F*_*i*_ and *R*_*i*_ with all other variables, and the third line follows from the mutual independence of the *D*_*ij*_ and from mutual independence of the *D*_*ij*_ and *S*_*i*_.

The appendix discussed the the bias arising from a selection rule that selects a line uniformly at random for each patient from the available post-platinum lines of therapy of that patient, in this general case where only *M*_*i*_ line starts are observed for each patient.

### Overall survival observed in retrospective database, choosing all eligible lines per patient

Instead of choosing a single eligible line for each patient, Hernán and Robins suggested that all eligible timepoints could be used. Applying this idea to our context, the comparison of external control arm patients to trial patients could be stratified by line, and an external control arm patient could be included in multiple strata, once for each line they had after platinum therapy. In each strata, the patient’s survival would be measured from the appropriate line of therapy and compared to patients enrolling in the clinical trial in that line setting. By equation 6, this would be an unbiased comparison, since the selection probabilities for patients in the external control arm would be *P* (*R*_*i*_ = *l* |*N*_*i*_ = *n*) = 1 regardless of the value of *n*. However, the correlations between survival times for patients included in multiple strata need to be accounted for. Here we account for these correlations using the grouped approximate jackknife estimate of the variance described in [10], which is implemented in standard statistical software [11].

### Simulation study

As shown above, if we choose a line of therapy at random for each patient in a retrospective database, but the random selection does not depend on *M*_*i*_, the number of lines observed for that patient, then our estimate of survival from each line setting will be unbiased. We can use this method to carry out a simulation in which we construct repeated single-arm trials, along with external control arms. Here the patients in both arms receive the same therapy and come from the same source population, so that if the external control arm is constructed correctly outcomes should be the same as in the single-arm trial.

To carry out this simulation study, we use the nationwide Flatiron Health electronic health record (EHR)-derived de-identified database. The Flatiron Health database is a longitudinal database, comprising de-identified patient-level structured and unstructured data, curated via technology-enabled abstraction. During the study period, the de-identified data originated from approximately 280 cancer clinics (800 sites of care). The study included 15,243 patients diagnosed with advanced non-small cell lung cancer from January 2011 to April 2020. The majority of patients in the database originate from community oncology settings; relative community/academic proportions may vary depending on study cohort. Lines of therapy in this database are oncologist-defined and rule-based. The data are de-identiifed and subject to obligations to prevent re-identification and protect patient confidentiality. Flatiron Health, Inc. did not participate in the analysis of this data.

We imagine we are using this dataset to construct an external control arm for a new compound being administered to lung cancer patients who have previously received platinum therapy. We allow patients in the external control arm to receive any therapy, i.e., we are comparing to physician’s choice of therapy, but we restrict to patients receiving therapy in the 5th or earlier line. There are 15,243 patients who meet these criteria in one or more line. 37% of these patients meet these criteria in only one treatment line; 30% meet them in two different lines and the remaining patients meet these criteria in three or four different lines.

We repeatedly simulate both a single-arm trial as well as an external control arm. Note that here the therapy is the same in the single-arm trial as in the external control arm, so that the survival distribution in both should be the same. To do this, for each simulation run we start by splitting our overall sample into two parts. From one part we select patients for the single-arm trial, using fixed probabilities to select patients in different line settings, so that on average trial patients will be evenly distributed across line numbers 2, 3, 4 and 5. From the other part we randomly select patients for the external control arm. We simulate single-arm trials and external control arms with 40, 160 and 640 patients.

We use three different methods to select lines for external control arm patients to compare to patients in the simulated single-arm trial: random selection of a line uniformly at random from the observed post-platinum lines of the patient, selection of the patient’s last line, and inclusion of all lines. We compare the three external control arms so constructed to the trial patients by calculating a hazard ratio (with the trial patients as the reference group), and its associated p-value, stratifying by line in a Cox proportional hazards model. We calculate bias as the mean hazard ratio (on the logarithmic scale) comparing the external control arm patients to the trial patients. In the absence of this bias this log hazard ratio should be equal to 0. We use 5,000 replicates per sample size.

Table 2 shows the bias that results from random selection of lines or using the last line for external control arm patients. This bias alone results in a hazard ratio of between 1.12 and 1.15 comparing external control arm patients to single-arm patients; this is an anti-conservative bias, as it will tend to exaggerate the effect of the therapy provided to single-arm patients. Table 2 also illustrates the lack of bias from the method in which all qualifying lines for all patients are used, with mean bias close to 0. Note that bias results from using random selection as the random selection of a line for an external control arm patient depends on how many lines a patient has in the database. Table 2 also shows the inflation of Type I error that results when using the naive variance estimator while including all lines for each external control arm patient; Type I error is more than doubled compared to the nominal 5% rate. For 640 patients, type I error is adequately controlled using the grouped approximate jackknife estimate, when all lines are used for each external control arm patients. Type I error is inflated only for lower sample sizes.

**Table 2:**
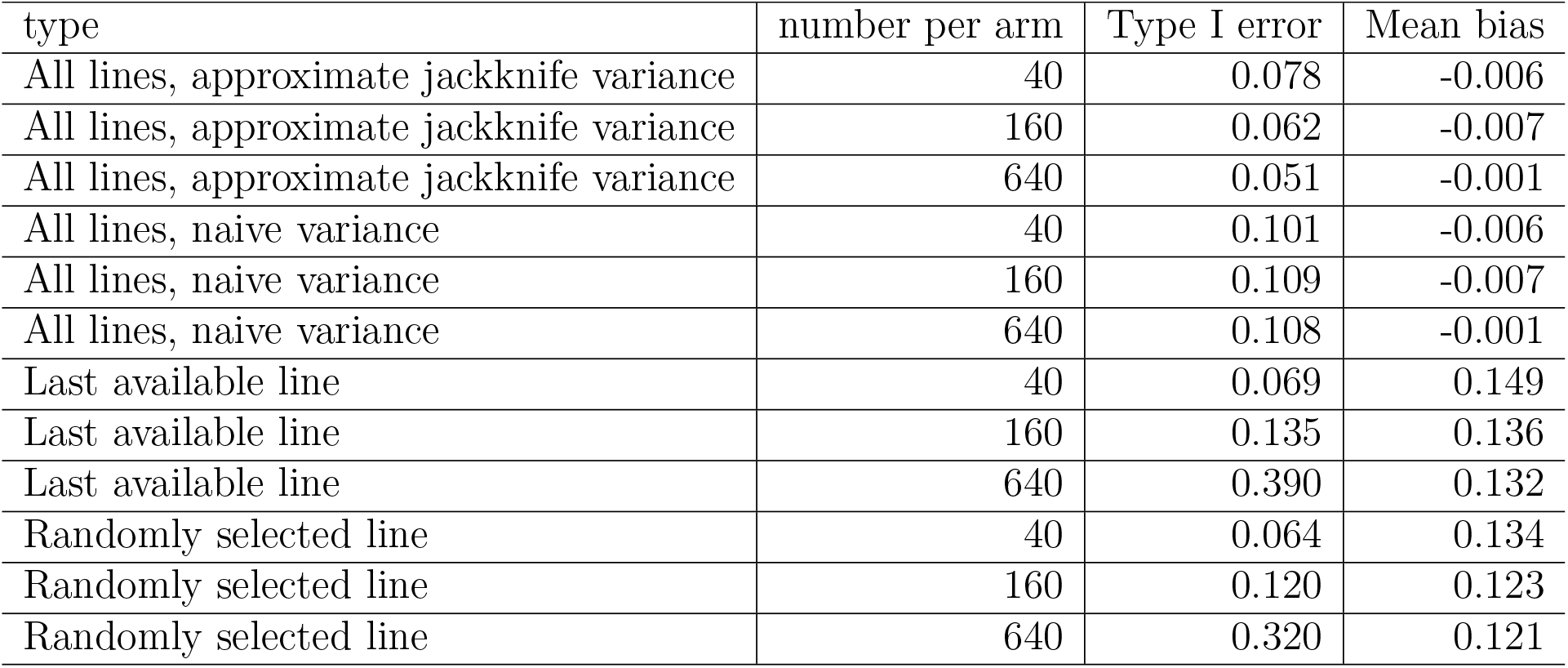
Simulation results. Mean bias is defined as the mean hazard ratio on the logarithmic scale, comparing simulated external control arm patients to simulated single-arm trial patients. In the absence of bias this log hazard ratio should equal 0. Testing was carried out at the 5% level so Type I error above 5% indicates inflation. For the all lines method, all available lines for external control arm patients were compared to the single-arm patients. For the last available line method, survival was measured for external control arm patients from the last line of therapy observed in the database. For the randomly selected line method, survival was measured for each external control arm patient from a line of therapy selected uniformly at random from among the available lines of therapy for that patient.

## Conclusion

What should be done when a patient in an external control arm for an oncology trial satisfies inclusion and exclusion criteria at the start of more than one line of therapy? Here we have shown that some methods of selecting a single line of therapy for each external control arm patient, like selecting a line of therapy uniformly at random, may lead to substantial bias, exaggerating the effect of the treatment provided to single-arm trial patients. These methods should therefore be avoided. By contrast, including all eligible lines for external control arm patients avoids this bias. We have demonstrated this via a model of the survival of cancer patients and their recruitment into a trial or inclusion in a retrospective database, as well as via a simulation study using data from a retrospective database. We have also demonstrated how to account for inflation of Type I error arising when more than one outcome for the same patient is included. The approximate jackknife estimator whose performance is evaluated here adequately controls Type I error, especially at larger sample sizes. Further work could assess whether the bootstrap better controls Type I error at smaller sample sizes.

## Data Availability

The data that support the findings of this study have been originated by Flatiron Health, Inc. These de-identified data may be made available upon request, and are subject to a license agreement with Flatiron Health; interested researchers should contact to determine licensing terms.

## 1 Appendix

### 1.1 Overall survival observed in clinical trial if *D*_*ij*_ are positively correlated

Assume that the *D*_*ij*_ are positively correlated even when conditioning on *N*_*i*_, the total number of lines of therapy, so that a patient that has one longer than average line of therapy is also likely to have other longer than average lines of therapy. As an example of the selection bias that can occur as a result, let us consider the probability that a patient who has *n* post-platinum lines of therapy will be selected into the trial at the start of their 2nd post-platinum line.

This probability is *P* (*S*_*i*_ *< t*_1_, *S*_*i*_ + *D*_*i*1_ *> t*_1_, *S*_*i*_ + *D*_*i*1_ *< t*_2_, *E*_*i*_ = 1|*N*_*i*_ = *n*). Let us assume that *S*_*i*_, the time of starting first post-platinum therapy, is uniformly distributed conditional on *N*_*i*_, i.e., that treatment patterns have not changed recently. Then this probability is proportional to *P* (*t*_1_ − *D*_*i*1_ *< S*_*i*_ *<* min(*t*_1_, *t*_2_ − *D*_*i*1_)|*N*_*i*_ = *n*) = min(*t*_1_, *t*_2_ − *D*_*i*1_) − (*t*_1_ − *D*_*i*1_). If *D*_*i*1_ *≥ t*_2_ − *t*_1_, the length of the enrollment period, then this expression is equal to a constant, *t*_2_ −*t*_1_. Otherwise, it equals *D*_*i*1_. We therefore see that the probability of selection in the trial is lower for patients with shorter *D*_*i*1_ than for patients with longer *D*_*i*1_. Since the *D*_*ij*_ are positively correlated, patients in the trial will then have longer post-enrollment survival than patients in the general population, conditioning on *N*_*i*_.

### 1.2 Selecting a line uniformly at random for each patient with censoring

When *N*_*i*_ is not observed for some patients in the retrospective database due to administrative censoring, then survival may be measured for each patient in the retrospective database from a post-platinum line of therapy chosen using a rule that depends on *M*_*i*_, the number of post-platinum lines of therapy observed for a patient in the registry. The survival for patients from line setting *l* in an external control arm so constructed is, conditioning on both *N*_*i*_ and *M*_*i*_ and applying Bayes’ rule:

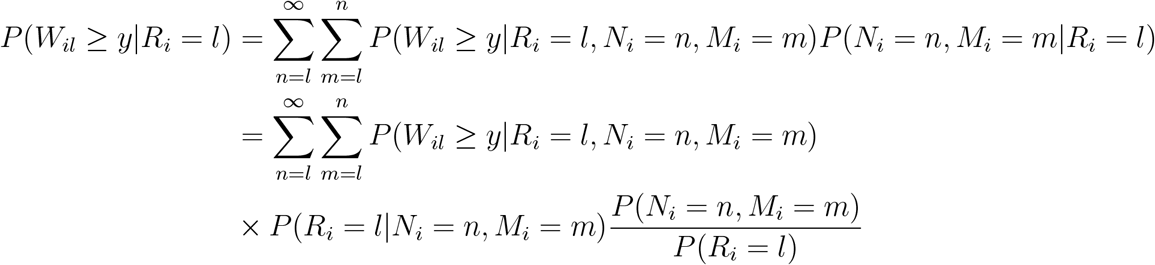

If we assume that a post-platinum line of therapy is chosen uniformly at random from among the *M*_*i*_ post-platinum lines of therapy for a patient, then this expression becomes

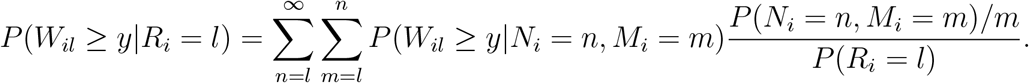

Here *W*_*il*_ is independent of *R*_*i*_ conditional on *M*_*i*_ since the selection of a line of therapy depends only on *M*_*i*_ and not on other features of the survival of the patient.

Let us assume, to keep things simple, that *N*_*i*_ can only equal 1 or 2, i.e., that patients can have at most 2 post-platinum lines of therapy. Had *N*_*i*_ been observed in this simple case and *R*_*i*_ had been uniformly selected from among 1, …, *N*_*i*_, then survival from the first post-platinum line for the external control arm would have been estimated (see equation 5) as

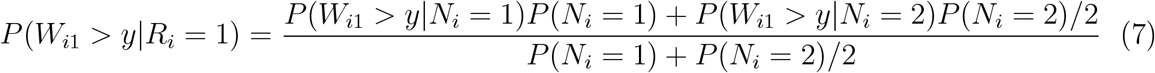

This is a biased estimate, compared to the survival probabilities as measured in a single arm trial (equation 2), due to the downweighting of the survival contribution from patients who actually survive for two lines of therapy.

In the general case where *N*_*i*_ is not observed due to administrative censoring, then instead survival from the first post-platinum line would be estimated as

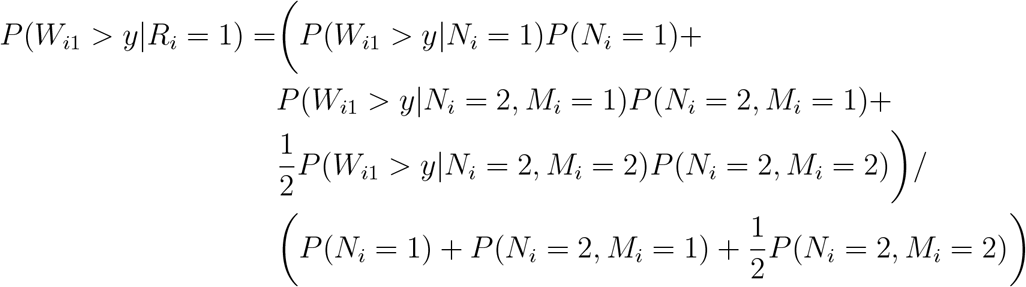

In this general case, we see that more weight is given in this weighted sum to survival from line 1 of those patients who actually survived for 2 lines, as compared to the biased estimate in equation 7. In particular, the contribution from patients who survive for 2 lines but whose start of line 2 was administratively censored is not downweighted at all. These patients may actually be expected to have better survival than those patients who survived for 2 lines but whose start of line 2 was not administratively censored, since patients with shorter first lines of therapy are less likely to be administratively censored. Therefore in this general case less bias is to be expected than in the case where *N*_*i*_ is observed for all patients.

Although bias arising from uniform random selection among available lines of therapy is attenuated as a result of administrative censoring, as compared to what would be expected from equation 5, it is important to note that most patients in a retrospective database may have observed dates of death, although the proportion will depend on how many years of data the retrospective database includes and the survival of patients in the database.

